# Quantifying the Risk of Indoor Drainage System in Multi-unit Apartment Building as a Transmission Route of SARS-CoV-2

**DOI:** 10.1101/2020.08.29.20184093

**Authors:** Kuang-Wei Shi, Yen-Hsiang Huang, Hunter Quon, Zi-Lu Ou-Yang, Chengwen Wang, Sunny C. Jiang

**Author notes:** Correspondence: Chengwen Wang, Room 735, School of Environment, Tsinghua University, Beijing, China, 100084, Sunny Jiang, 844E Engineering Tower, UC Irvine, California, 92697.

## Abstract

The COVID-19 pandemic has had a profound impact on human society. The isolation of SARS-CoV-2 from patients’ feces on human cell line raised concerns of possible transmission through human feces including exposure to aerosols generated by toilet flushing and through the indoor drainage system. Currently, routes of transmission, other than the close contact droplet transmission, are still not well understood. A quantitative microbial risk assessment was conducted to estimate the health risks associated with two aerosol exposure scenarios: 1) toilet flushing, and 2) faulty connection of a floor drain with the building’s main sewer pipe. SARS-CoV-2 data were collected from the emerging literature. The infectivity of the virus in feces was estimated based on a range of assumption between viral genome equivalence and infectious unit. The human exposure dose was calculated using Monte Carlo simulation of viral concentrations in aerosols under each scenario and human breathing rates. The probability of COVID-19 illness was generated using the dose-response model for SARS-CoV-1, a close relative of SARS-CoV-2, that was responsible for the SARS outbreak in 2003. The results indicate the median risks of developing COVID-19 for a single day exposure is 1.11 x 10^-10^ and 3.52 x 10^-11^ for toilet flushing and faulty drain scenario, respectively. The worst case scenario predicted the high end of COVID-19 risk for the toilet flushing scenario was 5.78 x 10^-4^ (at 95^th^ percentile). The infectious viral loads in human feces are the most sensitive input parameter and contribute significantly to model uncertainty.

## 1. Introduction

The ongoing COVID-19 pandemic^1^ caused by severe acute respiratory syndrome coronavirus 2 (SARS-CoV-2) has had a profound impact on human society and the world economy^2-3^. Despite the time and money invested in COVID-19 research and medical treatments around the world^4-8^, at the time of this writing, the diverse routes of transmission and effective treatment methods for this disease are still unclear. The fecal-oral transmission of SARS-CoV-2 has been a concern^9^ since the first detection of SARS-CoV-2 RNA from patients’ feces^10^. However, the infectivity of SARS-CoV-2 in feces and its hazard to human health is an ongoing debate although high concentrations of viral RNA have been found in both patient’s feces^11-13^ and in human sewage^14^. Only a handful of papers^10, 15-17^ reported the isolation of infectious viruses from feces on human tissue culture. The most recent report by Zang et al.^13^ provides evidence of SARS-CoV-2 replication in human small intestine but shows that most of the viruses are inactivated by simulated colonic fluid. This report explains the low frequency of viral isolation from fecal samples but it does not rule out the presence of infectious viruses in the patients’ feces as demonstrated by the other studies.^10, 15-17^ The rate of inactivation as the viruses passing through colon is likely dependent on time and protective effect of fecal material in colon. A fraction of viruses may survive the passage. Another contributor for the low frequency of fecal viral isolation on tissue culture may be due to the difficulties of measuring infectious virus in feces. Multiple purification steps are necessary to remove bacteria and other interferences before the samples can be loaded onto cell cultures. Such purification steps perhaps can inactivate or remove SARS-CoV-2. The methodological challenges can result in the uncertainties of the number of infectious virus in feces. However, based on what we know so far, the hazard of SARS-CoV-2 in feces seems likely to be real.

One of the possible transmission routes of fecal derived SARS-CoV-2 is through aerosols generated from toilet flushing and through indoor plumbing in multi-unit apartment buildings. The public concern was heightened by a report of high concentrations of SARS-CoV-2 virus in the aerosols of toilet rooms in two Wuhan hospitals^18^. Toilet flushing may release virus-laden aerosols and result in exposure^19-20^ of healthy individuals sharing the same bathroom. Furthermore, the commonly known sewer smell in bathrooms of multi-unit apartment building is suspected to be due to aerosols drawn in from the building’s main sewer pipe^19, 21^, which may contain viruses from neighbors’ toilets. The massive 2003 SARS outbreak in a condominium complex in Hong Kong, known as Amoy Gardens is still a fresh memory among many. The Amoy Gardens outbreak caused by SARS-CoV-1, a close relative of SARS-CoV-2, resulted in 321 cases of infection in the condominium complex. Investigations by the authority attributed the transmission to the indoor plumbing that drew contaminated aerosols from the patient’s toilet through the main sewer pipe connecting to different units in the same building^19, 22-23^.

In the last 20 years, researchers have investigated transmission risks from indoor plumbing and ventilation systems to provide recommendations for building construction and maintenance^19, 22^. Notably a number of studies have investigated the risk of *Legionella* transmission through indoor plumbing that delivers potable water or cooling water in multi-unit apartment buildings^24-26^. Building drainage systems in multi-unit apartment buildings are commonly served by a main sewer pipe, called a soil-stack (Fig. 1). Toilet flushing pushes human waste through the soil-pipe to the building’s main sewer soil-stack before leaving the building. However, viruses in feces can attach to the pipe wall and be present in sewer gas (aerosol) long after the waste has left the building. According to a recent laboratory decay study, SARS-CoV-2 can remain infective on metal and plastic surfaces for hours (the median half-life is around 13 hours on stainless steel and 16 hours on plastic surface) and has a median half-life of 2.7 hours in aerosol at 20°C^27^. The building’s main sewer soil-stack is connected to the floor drains and drains for sink, shower, and bathtub through a P-trap (Fig. 1). In normal situations, the P-trap retains a little bit of water after each use of the appliance. The water serves as a barrier to block the smell and aerosol from the main sewer pipe. When water in the P-trap evaporates, aerosols from the main sewer pipe can be drawn directly to individual bathrooms (commonly known as sewer smell in bathroom).

**Figure 1.**
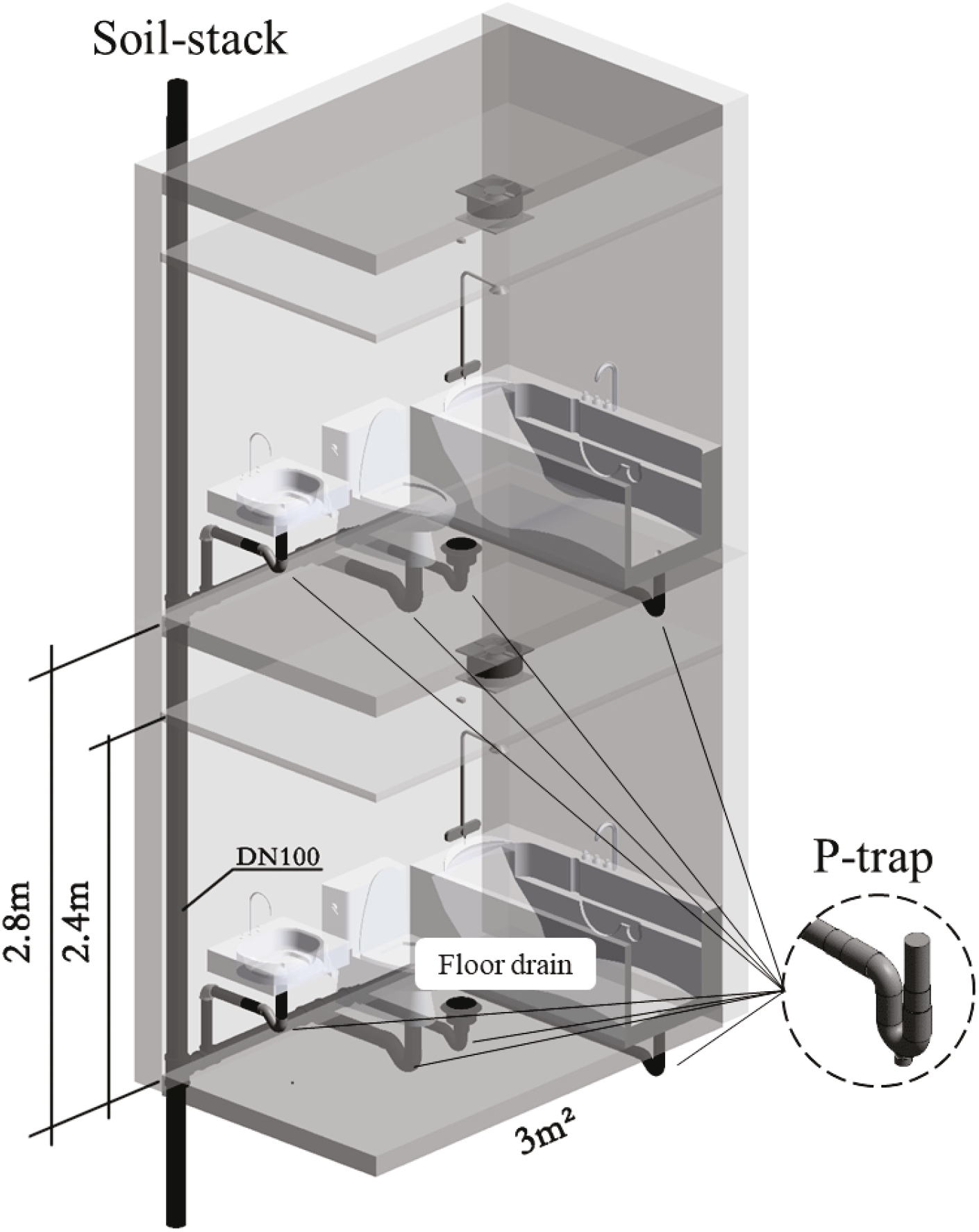
Illustration of indoor drainage in a multi-unit apartment building.

This study addresses the concerns of the aerosol generation from toilet flushing and their persistence in building drainage system^28-29^ using a quantitative microbial risk assessment (QMRA) for SARS-CoV-2 infection. Two possible transmission scenarios are analyzed: 1) inhalation of aerosols from toilet flushing of patient feces by a healthy person sharing the toilet room; and 2) inhalation of contaminated aerosols from the main sewer pipe entering the bathroom through a faulty floor drain by a neighboring resident. SARS-CoV-2 viral loads in human feces were collected from emerging literature reporting fecal viral loads^11-12, 16, 18, 30^. To compare and supplement the SARS-CoV-2 transmission model, SARS-CoV-1 data that caused the SARS outbreak in 2003 were also compiled from literature. SARS-CoV-2 exhibits long infectivity in aerosols, which is similar to SARS-CoV-1^27^. The similarities in genome sequences, human cell receptor for viral entry, and the environmental persistence between SARS-CoV-2 and SARS-CoV-1^27^ suggest that biological parameters for SARS-CoV-1 may be used as substitutes for SARS-CoV-2 in the absence of the SARS-CoV-2 specific data for risk quantification. In the absence of data on SARS-CoV-2, data on SARS-CoV-1 may be the best proxy.

## 2. Materials and Methods

### 2.1 Viral Concentration in Toilet Water and Aerosols

Many studies reported the detection of SARS-CoV-2 in COVID-19 patients’ feces^11-12, 16, 18, 30-33.^ At the time of this analysis, only a handful of the studies quantified viral load in human feces using qPCR^11-12, 16, 30 30^. Pan et al. reported 9 of 17 positive samples with a range of viral concentrations between 10^3^ and 10^5^ genome copies (gc) of viral RNA/mL of liquid stool sample (Table S1). The viral load varied over the course of the disease but was within 2 orders of magnitude. Wolfel et al.^11^ reported positive detection in 68 of 81 samples over 21 days for multiple COVID-19 patients. The SARS-CoV-2 concentrations ranged between 10^2^ and over 10^7^ gc/ gram of feces (Table S1). More recently, Zheng et al.^12^ followed patients for 60 days and detected 55 positive samples among 93 samples tested and reported the concentration in the range of less than 10^2^ and over 10^8^ gc/mL of feces (Table S1). Xiao et al.^16^ followed a single patient and quantified the viral load in feces by qPCR but used a standard curve based on known plaque forming unit (pfu). The viral load was expressed as pfu equivalence /mL of feces but indicated that fecal viral load does not necessarily imply infectivity.

The raw data reported in Pan et al.^30^ were provided to this study by the authors upon request. Individual data points from each of the other three reports were extracted from published figures using GetData Graph Digitizer^34^. In addition, a SARS-CoV-2 concentration of 19 gc/m^3^ of air measured directly in the aerosol from a hospital toilet room was also included in the analysis^18^. The data described above are compiled in Table S1 and the viral concentration in aerosols was assumed to be produced solely from flushing patients’ feces down the toilet.

To compare and contrast SARS-CoV-2 viral load in feces, we also searched and compiled data for SARS-CoV-1. As shown in Table S1, SARS-CoV-1 viral loads varied more significantly over the course of disease according to three different studies^35-37^. The SARS-CoV-1 concentration expanded over 6 orders of magnitude among different individuals and at different stages of disease.

Note that the viral concentration was reported as liquid volume of feces except in the study by Wolfel et al.^11^, in which viral RNA gc/gram was given (Table S1). Wolfel et al.^11^ indicated that the COVID-19 patients included in the study only had very mild symptoms and diarrhea was uncommon. To compile the data set, the values in per gram of feces were transformed to values in per mL using a fecal density of 0.97 g/mL^38^. The fecal viral concentration for both SARS-CoV-2 and SARS-CoV-1 were plotted in histograms and fitted with a cumulative density function curve shown in Fig. 2. The negative detections in each report were treated as below the lowest detection limit of 1.95 log_10_gc/mL by qPCR assay (Table S2). The values were generated by randomly sampling a uniform distribution U(0,1.95) for the fraction of negative detection reported (Fig. 2). Similarly, the below detection values for SARS-CoV-1 data were generated using U(0,0.35).

**Figure 2.**
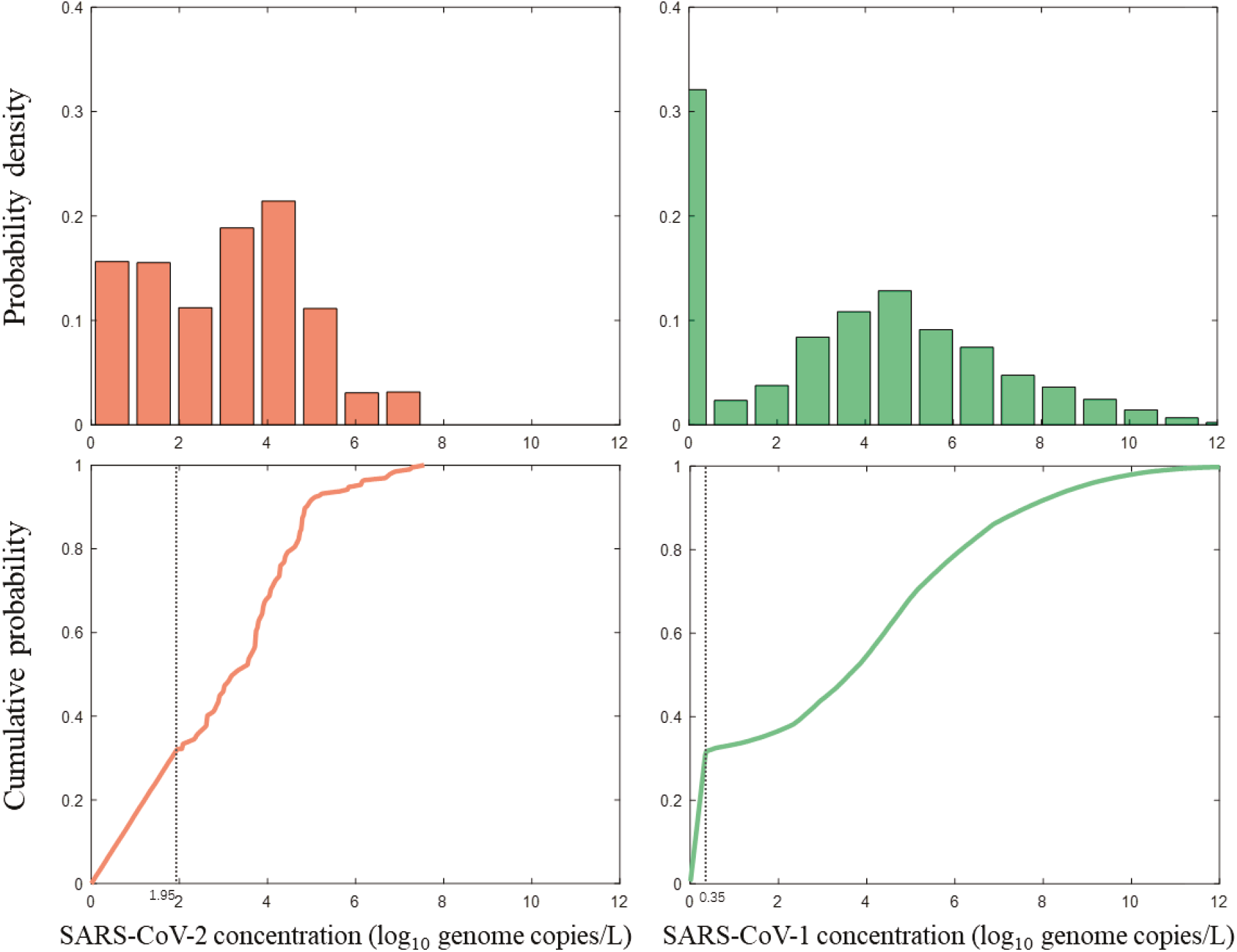
Histogram plots and cumulative density function curves of SARS-CoV-2 and SARS-CoV-1 viral loads in human feces (see Table S1 and S2 for data source). Note that the left side of cumulative distribution curve includes negative detection data from each report, which was presented as randomly generated data points using uniform distribution from zero to lower detection limit (doted vertical line).

The three data points from Xiao et al.^16^ was not included in the SARS-CoV-2 data distribution curve because they were expressed as pfu/mL. Instead a triangular(2.52, 2.97, 3.37) distribution was used to capture the range of viral load from this report (Table S2). The only direct toilet room aerosol measurement of 19 gc/m^3^ air was used as a single point estimate in risk analysis to represent an event as “aerosol measurement”.

An empirical distribution of human fecal volume (range 82 to 196mL, mean 135mL) per day^38^ and a uniform(3.78, 6.00) distribution of flushing water^39^ were used to estimate the concentration of virus in toilet water after defecation. The fraction of the infectious SARS-CoV-2 among reported genome equivalent viral RNA in feces is an important uncertainty. Previous studies of SARS-CoV-1 reported a wide range of conversion factor for viral genome number to infections unit (pfu), from 360:1 to 1600:1^40-41^. To include the uncertainty, a triangular(360, 980, 1600) distribution was used to capture the variability of this parameter (Table S2). Note that this conversion was not applied to the data reported by Xiao et al.^16^ since the fecal load was presented as pfu/mL. In the toilet, the patient’s stool was assumed to be completely mixed with flushing water. Thus, viral concentration in the flushing water after defecation (*C_fw,g,c,v_*, in gc/mL) is calculated using equation 1:

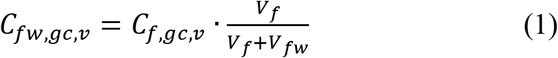

where *C_f_,_g,c,v_* is the concentration of virus in feces (gc/mL), *V_f_* is the volume of human feces per flush (mL), and *V_fw_* is the volume of flush water per flush (mL).

Viral concentration in the flushing water is then transformed into unit of pfu/L (*C_fw,pfu,v_*) by equation 2:

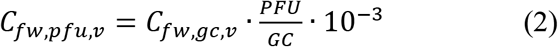

where 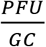 represents infectious units per viral gc (pfu/gc), and 10^−3^ is the unit conversion factor from per milliliter to per liter.

Aerosols generated from toilet flushing were assumed to contain the same viral concentration as the toilet water post defecation. The detailed data and assumptions are presented in Table S2.

### 2.2 Exposure Assessment for Aerosol Inhalation

Two exposure scenarios were considered to represent the generic living conditions in a multi-unit apartment building. The first scenario considers a shared bathroom by two suitemates in the same apartment suite. One resident is a COVID-19 patient under self-quarantine in his/her own room. The second scenario considers a COVID-19 patient under self-quarantine in an isolated apartment in the building. A neighbor in the apartment one floor below the patient’s apartment shares the main sewer soil-stack and has a floor drain missing water-seal to block the aerosol from the sewer pipe (Fig. 1).

#### 2.2.1 Toilet-flushing scenario

Under scenario 1, the aerosol concentration in the toilet room generated after a toilet flushing was adopted from a study by O’Toole et al.^42^. Since the aerosol concentrations were collected at different heights above the toilet in the original study, we adopted data collected at a sampling height of 420 mm above the toilet to represent aerosol concentrations inhaled by a person when using the toilet (Table S3). O’Toole et al.^42^ also showed a bimodal distribution of two median diameter aerosol sizes, d_1_=0.6 μm and d_2_=2.5 μm, at this height^42^. Although several other studies^43-44^ also reported the mass of aerosols generated by toilet flushing, the data were not comparable with the O’Toole et al.^42^ due to different types of toilet and different measurement approaches. These data were not included in aerosol estimation due to the need for assumptions of air volume and dispersion rate in the toilet room, which could induce additional uncertainty.

The decay of SARS-CoV-2 in the toilet-generated aerosols was computed using the half-life of 2.7 hours reported by van Doremalen et al.^27^. The healthy suitemate was assumed to be exposed to contaminated aerosols once a day between 0 and 2 hours after the patient flushed the fecal waste using U(0,2). The risk of exposure declines with the time after the prior flush due to viral decay in aerosols.

The aerosol deposition efficiencies in human respiratory tract were derived from Heyder’s study^45^, which was reported as a function of particle size and breathing patterns. The aerosol inhalation efficiency was based on an individuals’ breathing pattern during light activities^46^.

The duration of exposure for the healthy individual was set using a uniform distribution of 10 to 30 minutes U(10,30) to represent the various activities from urination to defecation and hand washing in the bathroom.

The dose of virus *(Dose_tf_,_v_*, in pfu/case) inhaled and deposited in the healthy resident’s respiratory tract under the toilet flushing scenario was estimated using equation 3:

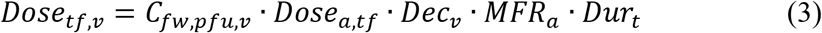

where *Dose_a,tf_* is the mass of aerosol according to median diameter size (*d_i_*) deposited in the exposed person’s respiratory tract (L/L of air), *Dec_v_* is the decay rate of virus in aerosols, *MFR_a_* is the mean flow rate of air breathed by the exposed person (L of air/min), and *Dur_t_* is the time spent in the toilet room each exposure event (min/case). The *Dose_a,t,f_* in equation 3 was derived using equation 4:

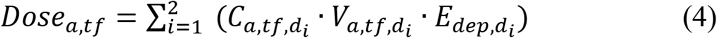

where 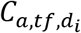 is the concentration of aerosols in the specific height above toilet after each toilet flush (# of aerosol/L of air) at range of diameter size *(d_i_*, in *μm)*, 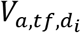 is the volume of spherical aerosol (L/# of aerosol), 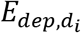 is the deposition efficiency of aerosols according to size in respiratory tract (unitless). The decay of virus in aerosols was calculated using equation 5:

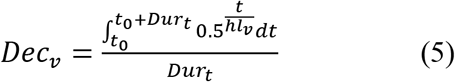

where *t*_0_ is the interval between the prior toilet flushing of patient’s feces and the healthy suitemate using the toilet room (min), and *hl_v_* is the half-life of the SARS-CoV-1 or SARS-CoV-2 in aerosol (min) according to van Doremalen et al.^27^. Parameters and assumptions used are summarized in Table S3.

#### 2.2.2 Faulty drain scenario

In this scenario, aerosols containing virus were assumed to be generated from the same flushing water and were suspending along the entire sewer soil-stack after the waste passed through the pipe. The aerosols in the sewer pipe were characterized once again by data from O’Toole’s study^42^ for samples collected immediately above the toilet (50 mm). A worst-case assumption was made that a ventilation fan drew all contaminated aerosols in the soil-stack pipe (0.1 m diameter and 2.8 m long between two floors) into the toilet room at the apartment one-story below through a floor drain that is missing the water-seal (Fig. 1). A toilet room size of 3 m^2^ x 2.4 m height, a typical toilet room in Amoy Gardens, was used to calculate the aerosol concentration. The dispersion of the aerosols in the toilet room was assumed to follow a Uniform(*log*_10_0.03,0) distribution.

Inhalation of polluted aerosols by the neighbor in the apartment below when using the toilet was modeled in the same way as the toilet flushing scenario. The dose of viruses through the faulty drain with no water seal *(Dose_fd,v_*, pfu/event) deposited in the exposed person’s respiratory tract was estimated using the equation 6:

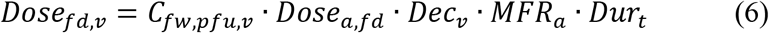

where *Dose_a,fd_* is the mass of aerosol according to median diameter size (*d_fd_*, *in μm)* deposited in the exposed person’s respiratory tract (L/L of air) in faulty drain scenario. It was calculated using equation 7:

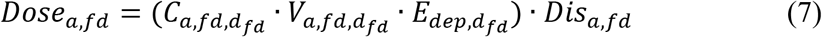

where 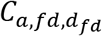 is the concentration of aerosols suspended in soil-stack pipe (# of aerosol/L of air), 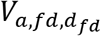 is the volume of spherical aerosol (L/# of aerosol), and *Dis_a,f,d_* is the dispersion rate of aerosol (unitless) in the toilet room. Parameters and assumptions used in this scenario are also summarized in Table S3.

The doses of exposure under different scenarios were compared. The simulated doses from fecal viral load were also compared with the exposure dose calculated using the SARS-CoV-2 concentration measured directly from the hospital toilet room^18^. Moreover, the model derived dose for SARS-CoV-1 under faulty drain scenario was compared to the dose estimated by Watanabe et al.^47^ in Amoy Gardens using a back calculation of attack rate and the dose-response model.

### 2.3 Dose-response Assessment

There has not been a dose-response model developed for SARS-CoV-2 for human or animals. SARS-CoV-2 shares 79% nucleic acid sequence identity to SARS-CoV-1, and uses the same cell entry receptor (ACE2) as SARS-CoV-1^48^. Structural analysis revealed SARS-CoV-2 protein binds ACE2 with 10-20 folds higher affinity than SARS-CoV-1, which indicates that SARS-CoV-2 may be more infectious to humans than SARS-CoV-1^48^. Moreover, SARS-CoV-2 triggers receptor dependent cell-cell fusion that helps the virus rapidly spread from cell-to-cell^49^. The basic reproductive values (R0) of COVID-19 at the early stage were calculated between 2 and 3.5, indicating that infected people on average could infect two to more than three other people, which was higher than SARS^50^. The effective reproduction number may be much lower due to less-susceptible hosts such as young adults^4, 51-52^. This infective pattern of SARS-CoV-2 is highly similar to SARS-CoV-1^53^. Based on the comparative biological property of the two viruses, we adopted SARS-CoV-1 dose-response model^47^ for SARS-CoV-2. To manage the uncertainty of this model, the probability distribution of *k* value in the exponential dose-response model was incorporated in the simulation to determine its impact on the uncertainty of model output. The normal distribution lnN(6.01,1.75) as originally reported for SARS-CoV-1 dose-response by Watanabe et al.^47^ was used in the simulation. Moreover, the *k* was customized to SARS-CoV-2 by including the enhanced infectivity factor using uniform distribution U(10, 20) to capture the 10 to 20 times higher cell-receptor affinity and viral spread through cells. Since it is currently unclear that the ACE2 receptor binding affinity would linearly translate to infectivity in doses, this general model was subjected to sensitivity analysis. Therefore, the same dose-response model (equation 8) for SARS-CoV-1 and SARS-CoV-2 was used with the different best fit *k* value.

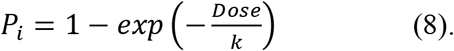

The parameters used in the dose-response assessments are presented in (Table S4).

### 2.4 Risk Characterization

To estimate the risk of developing COVID-19 by the suitemate or the downstairs neighbor, we assume a once-a-day encounter rate of polluted aerosols over a 15-day course of disease (Table S4). The estimations were carried out using equation 9 and the illness rate for COVID-19 was compared to the simulated outcomes for SARS, to provide a relative risk perspective.

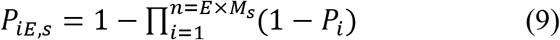

Where *E* is the extension of the course of COVID-19 assuming 15 days, *M_s_* is the frequency of a scenario occurring in a day.

Moreover, the aerosol concentration measured directly in the toilet room used by multiple COVID-19 patients in a Wuhan hospital of 19 genome copies/m^3^ air^18^ was included in the risk estimation to represent the worst case scenario of COVID-19 transmission. The risk estimation using data from Xiao et al.^16^, where the fecal viral shedding was presented as pfu/mL, was presented separately.

### 2.5 Monte Carlo Simulation and Sensitivity Analysis

The pseudo-algorithm information flow is shown in Figure S1. The input parameters were randomly sampled from their established probability distributions. 100,000 iterations of each output parameter were computed to ensure the distributions reach a steady state. Reproducibility of the outputs is examined by a variation of less than 1%^54^. All computations were carried using MATLAB R2019b^55^.

A sensitivity analysis was conducted to determine which model inputs were the most influential contributors to the predicted illness risk. The rank of importance was introduced through incorporation of parameter sensitivity with the relative order of uncertainty to assess the confidence in the model. The importance factor (*I*) contributing to illness risk for each input parameter (unitless) was calculated using equation 10:

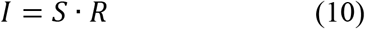

where *S* is the sensitivity of illness risk related to the input parameter, and *R* is the coefficient of variation of the input parameter.

*S* was assessed by a local sensitivity analysis method to represent the variability propagation of input parameters through modeling of the health risk. The true means of distributions (or the values of point-estimates) were adopted as baseline point values for each input parameter and output variable. Then the baseline input parameter *P_m_* value was decreased by 10%, and a differential value for output variable *X_m_* was calculated as shown in equation 11:

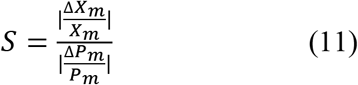

where *X_m_* is the mean value of the original output variable distribution, Δ*X_m_* is the difference in means between the original output distribution and the altered output distribution, *P_m_* is the original baseline point value, and Δ*P_m_* is the difference between the original baseline value and the altered value.

*R* was incorporated to represent the uncertainty of input parameter distribution, and was calculated using equation 12:

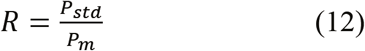

where *P_std_* is the standard deviation of the input parameter.

## 3. Results and discussion

### 3.1 Comparison of Exposure Dose Under Different Scenarios

The exposure doses of SARS-CoV-1 and SARS-CoV-2 per event under different scenarios are presented in Fig. 3. The simulated doses based on fecal load had a bimodal distribution because of the inclusion of non-detectable results. The median values for SARS-CoV-1 and 2 were similar, in the range of 1.15 to 8.45 x 10^-9^ pfu per exposure event (see Table S5 for summary descriptors). However, comparing the two viruses, SARS-CoV-1 had a wide range of exposure dose with a long flat tail (95^th^ percentile is 1.58 x 10^-3^ pfu for toilet flushing scenario) on the right side of the distribution curve (Fig. 3 and Table S5).

**Figure 3.**
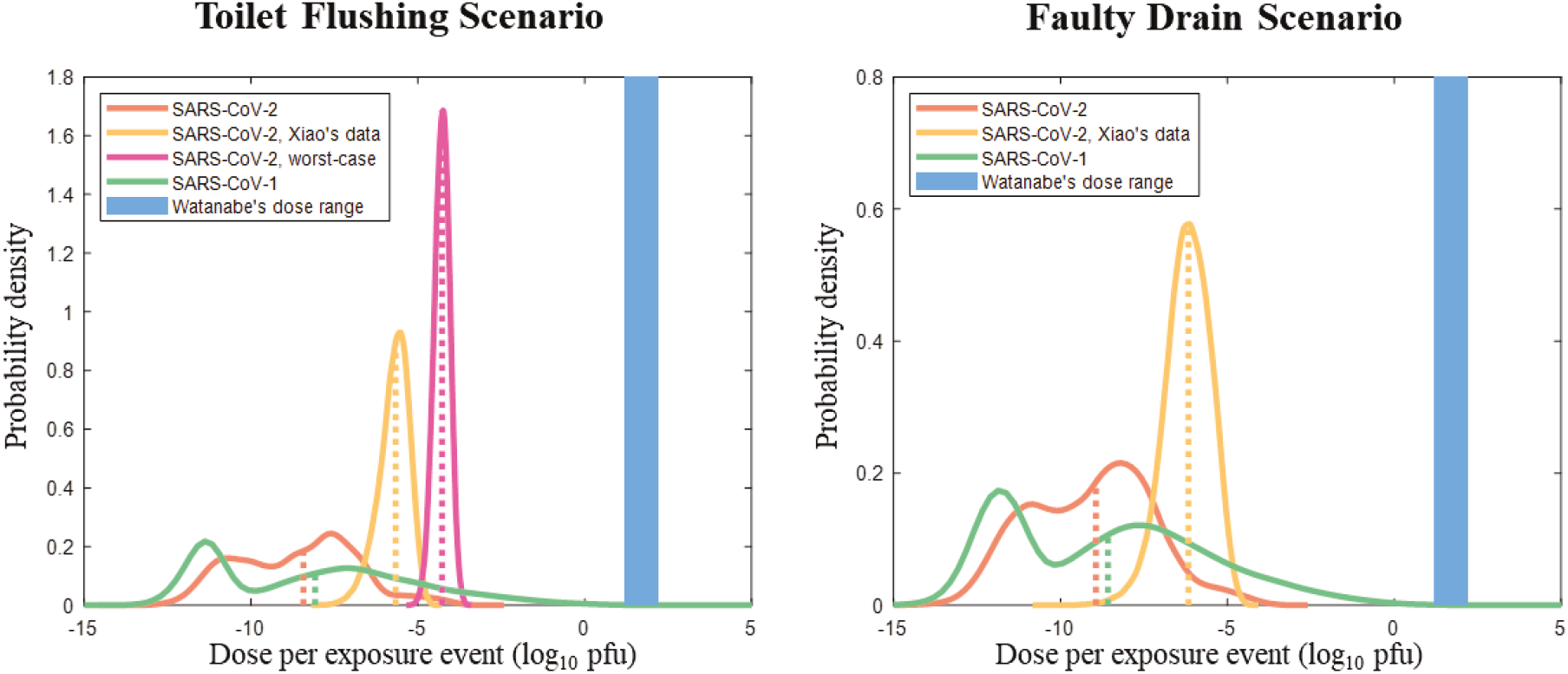
Distribution plots showing doses of SARS-CoV-2 and SARS-CoV-1 per exposure event, respectively, estimated using different scenarios and data source. SARS-CoV-2 worst-case is from the aerosol measurement from Wuhan hospital toilet room. Dotted lines represent the medians of distributions. The vertical blue bar is the range of doses estimated by Watanabe et al. for Amoy Gardens’ incident.

Using fecal load data reported by Xiao et al.^16^ (in pfu/mL) to calculate the exposure dose shifted the median exposure dose to the right by over 2 to 3 log10 units (Fig. 3, Table S5). However, this exposure dose was likely an overestimation of infectious SARS-CoV-2 because the authors of the fecal shedding study indicated that the pfu equivalent numbers reported do not indicate infectivity number^16^. It is important to note that the pfu equivalent presented in Xiao et al. is not the same as the viral infectious unit presented elsewhere in the study. The worst case scenario of exposure to SARS-CoV-2 was represented by exposure to an aerosol concentration measured directly in the hospital’s toilet room (Fig. 3). The median exposure dose of this scenario was 5.34 x 10^-5^ pfu per exposure event (Table S5).

There is a dramatic mismatch when comparing the exposure dose estimated based on any of the scenario with the exposure dose calculated by Watanabe et al.^47^. Watanabe et al. based their estimaion on the attack rate in Amoy Gardens and the dose-response relationship derived using an animal model (Figure 3). The explanations for the mismatch are complicated and may include the following reasons.

1. Watanabe et al.^47^ assumed that all infections in Amoy Gardens were resulting from the faulty drainage system in the building and calculated the attack rate based on estimated number of residents per unit. This attack rate may be an overestimation because other modes of transmission (i.e. surface contact and in person transmission in shared elevators or space) besides aerosols through faulty drains were not considered^56-57^.
2. Our aerosol concentrations are underestimated by several orders of magnitude. Since the initial infectious viral load (pfu) in feces is a sensitive input parameter that influences the aerosol dose in the toilet room, the major uncertainty could be from the conversion of viral genome copy to infectious unit. We used SARS-CoV-1 as the surrogate to get the conversion factor from laboratory studies by different researchers^40-41^. We used triangular distribution to derive the range. The outcomes of the conversion were ~100 times lower than the report presented by Xiao et al.^16^ who used pfu equivalent directly as the standard curve in the estimation of viral load in feces. Another source of underestimation is from the literature reports of fecal loading among COVID-19 patients. The data collected in this study include a total of 191 human fecal samples (exclude the 3 samples form Xiao et al.^16^) at different stage of disease development. It represents the state of knowledge at the time of this analysis.
3. The dose-response model derived from animal study and used by Watanabe et al.^47^ in model fitting is an overestimation of the infectious dose of the SARS-CoV-1. Based on the animal trial study^58-59^, the SARS-CoV-1 requires higher dose of infectious than HCoV-229E, the common human cold virus. The SARS-CoV-1 dose-response was investigated using genetically modified mice, which may not be a good representation of human infection rate.

Based on these above analyses, the unmatched dose in aerosol exposure between Watanabe et al.^47^ and our model prediction cannot be easily resolved. The uncertainty analysis that incorporates the sensitivity with the coefficient of variation of the input parameter (as presented in the section 3.3) weighs the importance of the input parameters to the model outcomes. This first attempt at the risk analysis of the specific scenario offers a starting point for looking into data collection needs.

### 3.2. Risk Quantification Per Exposure Event and Per Course of Disease

Fig. 4 shows the outcomes of risk estimation for different scenarios. The median COVID-19 illness risk for a single day exposure is 1.11 x 10^-10^ and 3.52 x 10^-11^ for toilet flushing (Fig 4a and Table S6a) and faulty drain (Fig 4b and Table S6a) scenario, respectively. These values are nearly one log higher in comparison with the SARS illness risk predicted using SARS-CoV-1 fecal loading estimation, which have a median value of 1.75 x 10^-11^ for toilet flushing and 5.5 x 10^-12^ for the faulty drain scenario (Table S6a). However, there is a large variability of predicted risk for SARS with the 95^th^ percentile risk of 5.28 x 10^-6^ and 1.82 x 10^-6^ for toilet flushing and faulty drain scenario, respectively. The corresponding 95^th^ percentile risks for COVID-19 are 1 to 1.5 log lower.

**Figure 4.**
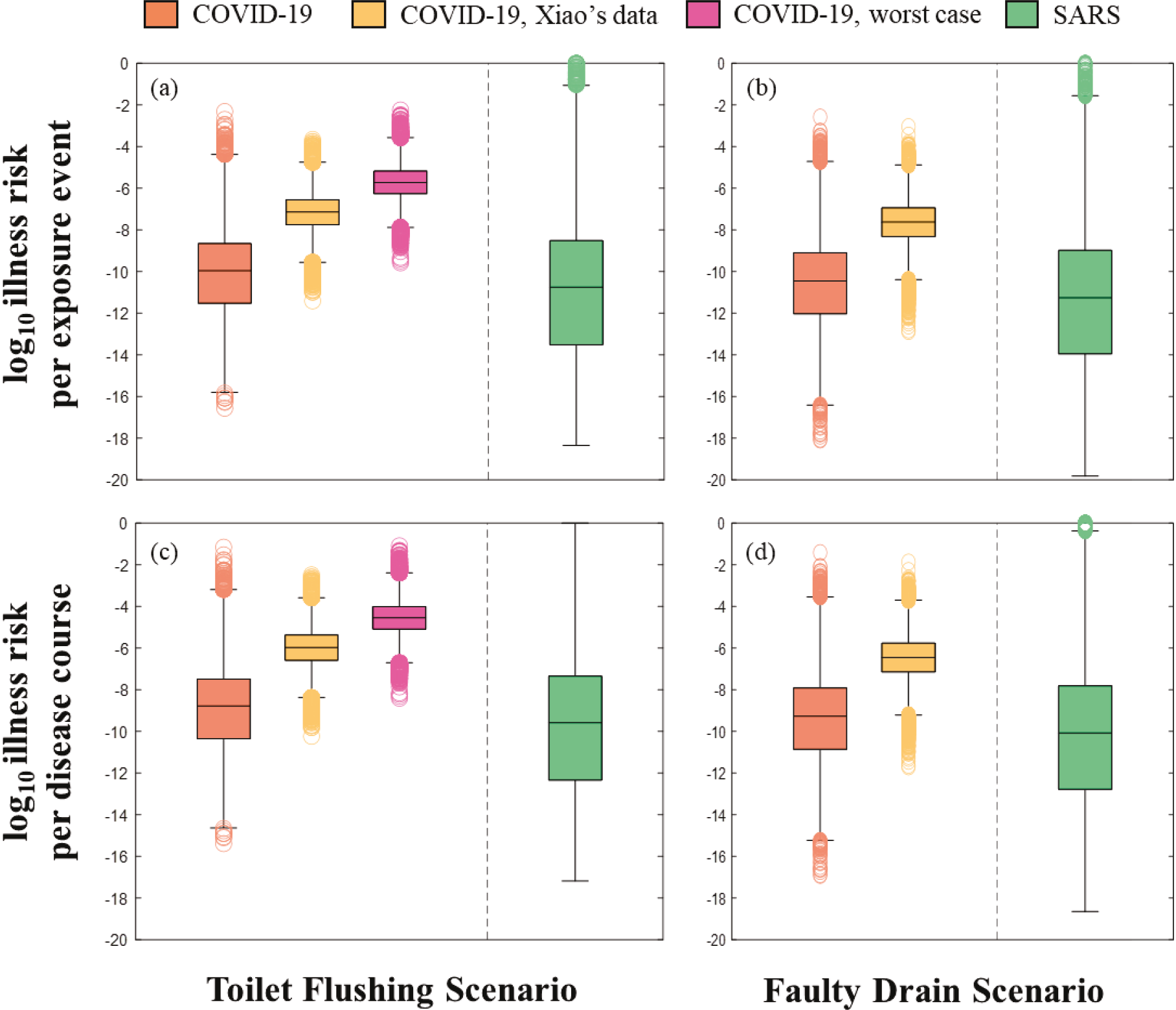
Box-and-Whiskers plots showing COVID-19 and SARS illness risks per exposure event (a & b) and per disease course (c & d) using data collected for SARS-CoV-2 and SARS-CoV-1, respectively. Each box represents the 25^th^, median (50^th^), and 75^th^ percentile values of the distribution, where the whiskers extend 1.5 x (75^th^ percentile value - 25^th^ percentile value) from each end of the box.

Xiao et al.’s fecal load data^16^ predicted much higher risks for both scenarios with the median risk in the order of 10^-8^ and 95^th^ percentile risk in the order of 10^-6^ (Fig. 4 and Table S6a). The estimates for exposure to contaminated aerosols in the hospital toilet room yielded a median risk value of 1.9 x 10^-6^ and 95^th^ percentile risk of 3.85 x 10^-5^ for each single exposure (Table 6a).

When multiple exposures were considered over the 15-day course of the disease, assuming once a day exposure frequency, the estimated risk of COVID-19 increased by approximately one log for all scenarios (Fig 4c,d and Table S6b). Again, these values were about one log higher than median illness risks for SARS under the same scenarios but SARS outcomes spanned a much greater range (Fig. 4 c, d and Table 6S). The worst case scenario, exposure to aerosol in a hospital toilet room used by multiple patients, predicted the high end of COVID-19 risk for toilet flushing scenario at 5.78 x 10”^4^ (95^th^ percentile). Comparing risks from the toilet flushing with the faulty drain scenario, it’s obvious that direct exposure to aerosols generated from toilet flushing had greater risk than exposure to aerosols entered from a faulty drain in the building (Fig. 4).

These risk estimates are at odds with the conclusion of Amoy Gardens’ investigation, where the epidemiological study presented a very different picture, a much higher attack rate from the faulty drain scenario. The uncertainties of a number of input parameters in the model are worthy of analysis to offer better insights into the discrepancies observed from the two investigations. These input parameters include the concentration of viruses in the feces, the conversion factor from genome equivalent of virus to infectious unit, the daily fecal volume, the toilet flushing water volume, the aerosols generated during toilet flushing, the duration of exposure in the toilet room, the viral decay and the best fit parameter for dose-response model.

### 3.3 Sensitivity Analysis and Model Uncertainties

The analyses for model uncertainties were assessed using the joint estimation of local sensitivity of each parameter and the coefficient of variant for the parameter to evaluate the relative order of uncertainty. Note that the analysis was only carried out for models starting with genome equivalent SARS-CoV-2 fecal loadings. The single aerosol measurement from the hospital toilet room was not a good representation of the normal living condition in the multi-unit apartment building and only presented here as the worst case scenario. The results showed that the concentration of genome equivalent virus in toilet flushing water *(C_fw,gc,v2_)* was the most important input parameter in the risk estimation for both scenarios (Fig. 5). Variability of this parameter contributed to 48% of the variability of model outcome and represented the most uncertainty. The conversion factor (PFU/GC) that bridges the genome copies of virus to pfu of the infectious virus, in comparison, was only a minor contributor (1 to 2% of variability) to the overall model outcome (Fig. 5). The concentration of aerosols in the toilet room (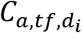 and 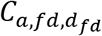) had a more important role than the genome conversion factor and attributed to 4 and 5% of the variability in the model output. Unique to the faulty drain scenario, the aerosol dispersion rate *(Dis_a,fd_*) from the drain to the toilet room was a sensitive and uncertain parameter, which contributed to 7% of the variability in the risk outcome. The fecal volume (*V_f_*), time spent in the toilet room *(Dur_t_*), the SARS-CoV-2 decay half life *(hl_v2_)*, and time delay (*t*_0_) after the prior toilet flushing all contributed to 2% of the variability in the model outcome and were less important in comparison with the concentration of the virus in the flush water (*C_fw,gc,v_*_2_). Aside from the factors influencing the exposure risk, the dose-response best fit parameter *k_v2_* in the exponential dose-response model was the second most important contributor to the variability of the illness risk estimation. 37% of variability in illness estimation was due to this parameter in the toilet flushing scenario and 34% in the faulty drain scenario.

**Figure 5.**
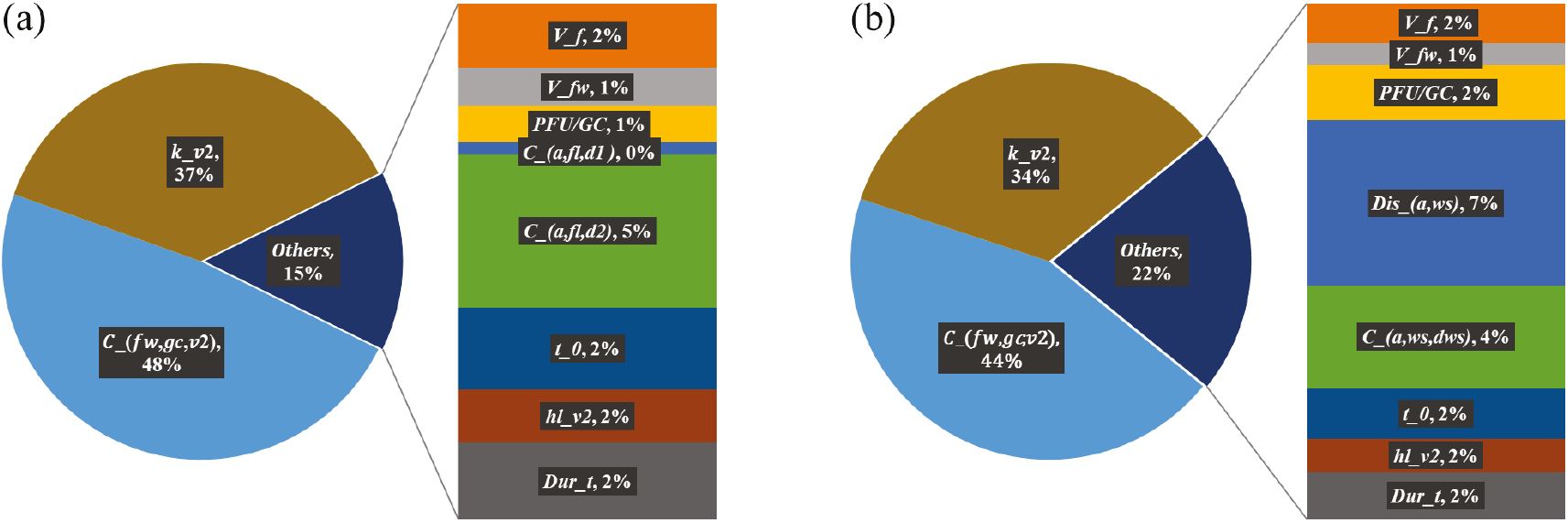
Ranked importance analysis of model outcomes. See manuscript for parameter descriptions.

We initially expected the conversion factor from genome copy virus to infectious virus pfu was a major source of model uncertainty since determining the concentration of infectious SARS-CoV-2 in the fecal samples had been challenging^11^. The above analyses showed, however, the wide range of the viral load in the fecal shedding played an important role. In the effort of the data collection, we focused on literature that provided quantitative analysis of fecal loading in patients at different stage of the disease development and from different regions. We also included data reported as non-detection by setting the lower detection limit as the upper bound to generate estimations using a uniform distribution between zero and the detection limit. The SARS-CoV-2 data compiled here represent the state of knowledge and are comparable with SARS-CoV-1 data, which served as a proxy for the new virus.

To further assess the confidence of fecal viral load among COVID-19 patients, we compared the estimated total viral load from all COVID-19 cases (15-day accumulated cases) in a sewage treatment plant’s service area with the reported concentration of SARS-CoV-2 in sewage. The back-of-the-envelope calculation was carried out using the City of New York COVID-19 cases reported during their peak outbreak. The case numbers were collected in Newtown Creek Wastewater Treatment Plant (NCWTP) service area, including Manhattan, Brooklyn and Queens^60^. The case numbers were multiplied by the fecal loading rate and divided by the influent volume of raw sewage to NCWTP. The details of the calculation and outcomes are presented in the supporting materials and Figure S2. The analysis showed that based on reported cases and fecal viral load, the median concentration of SARS-CoV-2 was estimated to be 1.25log_10_gc/L of raw sewage without including viral decay during transport from households to NCWTP (Table S7). If assuming reported cases were only 50% of the total number of cases in the community (due to underreporting, asymptomatic and pre-symptomatic cases), the estimated concentration of SARS-CoV-2 would only increase to 1.55log_10_gc/L. Even assuming 50% of all residents in the plant service area were COVID-19 patients and shed virus in the feces, the median concentration of SARS-CoV-2 was still in the range of 3.1log_10_gc/L of raw sewage. In comparison with the reports of SARS-CoV-2 concentration in sewage from different municipal sewage treatment plants (median value of 5.23log_10_gc/L), the estimates based on fecal loading were much lower (Figure S2 and Table S7). This result suggests that the fecal loading is either underestimated or sewage SARS-CoV-2 concentration is several orders overestimated. The underestimation of fecal viral load could be due to the methodological challenges relating to sample extraction and purification of viruses for qPCR. Since the uncertainty of this parameter plays a key role in the risk estimation, future data in this area will no doubt improve the confidence in the risk estimation.

Regardless of the contribution of the conversion factor to model variability, the infectivity of the viruses shed in feces is a critical factor to establish the hazard in discussion. At the time of this writing, there are two sides of the expert opinions regarding the infectivity of SARS-CoV-2 in feces. One side believes that viruses shed in feces are mostly inactivated by colonic fluid^13^, therefore, there is low or no hazard from fecal shed virus. So far, there has not been any direct evidence to support fecal transmitted infection in human or in animal model for either SARS-CoV-2 or SARS-CoV-1. The conclusion from the Amory Gardens investigation was challenged by previous reports^56-57^. The other side of the opinion points to the successful isolation of infectious virus from fecal samples from the handful of presence and absence studies to support the hypothesis of fecal-oral transmission^10, 15-17^ Several reports also indicate that viral shedding in feces long after patients recovered from COVID-19 and tested negative for SARS-CoV-2 in respiratory samples. They suggested that prolonged viral shedding could pose repeated exposure risks to others even after the patient has recovered from disease^32^. In addition to the case report of Amoy Gardens for SARS-CoV-1 transmission, two recent governmental reports from China concluded that fecal exposure was the likely source of COVID-19 transmission. One was a press release of the Infectious Disease Control Authority in the City of Guangzhou, China, which pin-pointed the COVID-19 patient feces as the source of infection of three families residing in the floor below a COVID-19 patient ^61^. The second was a report by Beijing Center for Disease Control and Prevention identifying that an isolated incident of a COVID-19 case was linked to a known cluster of outbreak through a visit to a public bathroom^62^. More research is needed to understand infectious potential of feces.

We adopted the genome copy to infectious unit conversion of SARS-CoV-1 reported in the lab experiments as a proxy for SARS-CoV-2. It is important to point out that the SARS-CoV-1 used in these experiments was not derived from fecal samples^40-41^. They were lab viral cultures, and the relationship between genome copies and infectious units depended on the methods for virus preparation, the storage condition and time delay before the assay. If we believe the colonic fluid can effectively inactivate SARS-CoV-2, the conversation factor adopted from lab cultured viruses could be an overestimation of infectious virus in feces and the final risks. The conversion factor used in the model covers a range based on the lab experimental outcome but it may not be the right representation of fecal shed virus. The degree of uncertainty in this factor may not be adequately represented by the importance analysis. Until further data on infectivity of SARS-CoV-2 in feces is made available, this is the best proxy for converting genome equivalent to infectious unit.

The aerosol generation during toilet flushing has been well studied^20,21^. Aerosol concentration varies with types of toilet. The aerosol concentrations presented by O’Toole’s study^42^ are the best representation of the low flow toilet commonly installed in the multi-unit apartment buildings. The importance analyses indicated both aerosol concentration and dispersion in a room could have important impact on the risk outcomes. However, they were unlikely to change the magnitude of the risk that could match the risk outcome of Amoy Gardens. In comparison with the direct measurements of aerosols generated by toilet flushing, the dispersion of aerosols through a faulty drains is less certain. The commonly known sewer smell only provides indirect evidence for aerosol intake from the main sewer pipe to individual toilet rooms. We built a uniform distribution for the dispersion rate to capture the variability of this parameter. Additional research is needed to understand the building drainage system and aerosol transmission.

Human dose-response model for SARS-CoV-2 requires intentionally exposing humans to known doses of SARS-CoV-2, which is unlikely due to the ethical concerns. Animal models may be developed in the future as that has been done for SARS-CoV-1. Although there are remarkable differences between SARS-CoV-2 and SARS-CoV-1 during infection, the disease development shares similarity, which promotes us to use the SARS-CoV-1 model as the proxy. Analyses showed that this was a critical parameter of the risk model and contributed to a large degree of uncertainty in the risk outcome. Adjustment made in the *k* through incorporation of enhanced affinity to cell receptor, did not dramatically increase the risk. The uncertainty was largely embedded in the original model parameter, where data were pooled from both the animal model and comparison with other coronavirus including HCoV-229E to generate the best fit *k*. Together with viral load, *k* had the biggest impact on risk outcomes. However, even after removing the uncertainty of the dose-response relationship, the comparison of the exposure dose estimated by the model still could not resolve the large gap with the dose estimation from Watanabe et al.^47^. Therefore, this analysis suggests additional routes of transmission may have contributed to Amoy Gardens outbreak^57^.

### 3.4 Implications and Recommendations

The risk estimation presented relatively low median health risks of two viral transmission scenarios through building drainage system. However, there are large degrees of uncertainty among several model input parameters. Although uncertainties are inevitable, risk assessment should always be conducted in an iterative manner that allows refinement of the risk assessment question(s), key assumptions, and data used in the model. As the first attempt to understand the risk of a novel virus, we expect the risk analysis to offer the fundamental understanding of associated risk based on the risk analysis framework. The model and outcomes can be refined in the later time with the emergence of new data on the property of the virus, human and environmental interactions.

The mismatch of the exposure dose and illness estimation of this study with Amoy Gardens’ report is worthy of further investigation. Although the high end of the risk estimation does not exclude the transmission scenario, it has a lower possibility or is only under extreme conditions. The extreme conditions may include a partially stopped up drainage system that can trap human waste containing virus in the sewer pipe, overloading ventilating fan to draw contaminated aerosols constantly into the toilet room^19^. Therefore, attentions should be given to proper maintenance of building drainage systems during the outbreak of aerosol transmitted diseases.

It should be noted that the existing indoor drainage systems, if used and maintained properly, are able to protect healthy habitants from the possible exposures to pathogens, which is validated by the lack of more incidents like Amoy Gardens. Recommendations such as keeping the toilet cover closed while flushing to prevent the aerosols from being splashed into the air, and a regular inspection of water seals to prevent the aerosols in drainage pipes from being released into the indoor air could be made to further reduce the risk. Such simple habits could effectively keep habitants from microbial risks associated with the building drainage system.

The toilet flushing and faulty floor drain scenarios are examples of potential hazards of the fecal contamination derived risk. The aerosolization of fecal derived virus can also be applied to risk analysis in shared public restrooms since the restroom can be filled with aerosols generated by multiple toilet flushes within a short time window. The viral concentration in the air could be significantly higher if there were large numbers of COVID-19 patients or asymptomatic carriers using the restroom (maybe closer to the direct measurements in the Wuhan Hospital toilet room). Another potential application is to look into the aerosol generation from aerated sewage during wastewater treatment process to estimate the risk of personnel working in wastewater treatment plants.

## Data Availability

Data are available upon request

## Acknowledgments

Funding for this research is provided by National Science Foundation grant CBET 2027306 and UC Irvine COVID-19 CRAFT research fund to S. Jiang and by Chinese Major Science and Technology Program for Water Pollution Control and Treatment (No. 2018ZX07110-008) to C. Wang. The authors acknowledge the contribution of original data for fecal viral loads by Dr. Poon (Pan et al., 2020) and feedback from Y. Ren and H. Wang during the project development.

## Supporting Information/Data in Brief

Data for SARS-CoV-2 and SARS-CoV-1 in human feces; parameters used in hazard identification, exposure assessment, dose-response and risk characterization; pseudo-algorithm flow chart for estimating health risks; summary descriptors for dose estimation, and risk per exposure event and per disease course; comparison of total fecal derived SARS-CoV-2 load in sewage with SARS-CoV-2 measured directly in sewage.

## CRediT Statement

Kuang-wei Shi: data collection, analysis and writing; Yen-Hsiang Huang: data collection and analysis; Hunter Quon: data collection and editing; Zi-Lu Ou-Yang: data collection; Chenwei Wang: conceptualization; Sunny Jiang: conceptualization, data collection, writing, review and editing.

## Notes

### Competing Interest Statement

The authors have declared no competing interest.

